# Motor Unit Number Index (MUNIX) of the Upper Trapezius: Reliability and Meta-Analysis

**DOI:** 10.1101/2021.03.14.21253565

**Authors:** Agessandro Abrahao, Liane Phung, David Fam, Marcio Luiz Escorcio-Bezerra, Lawrence R Robinson, Kelvin E Jones, Lorne Zinman

## Abstract

Motor unit number index (MUNIX) of the upper trapezius is a candidate biomarker for lower motor neuron function of the bulbar region; however, only a few studies have explored this measure in neuromuscular diseases and reliability data is incomplete. We conducted a systematic review and meta-analysis of this measure in control participants and assessed its reliability in twenty healthy volunteers. Four studies were included with heterogeneous mean-MUNIX estimates, moderated by variability in the population’s age and MUNIX sampling technique. We demonstrated an inter- and intra-rater intraclass correlation of 0.86 and 0.94, respectively. Upper trapezius MUNIX is a reliable measure with in-between study variability moderated by age and MUNIX technique.

## Introduction

Motor Unit Number Index (MUNIX) is a neurophysiological technique that indirectly estimates the number of functioning motor units of a given muscle. MUNIX uses the compound motor action potential (CMAP) and surface interference pattern (SIP) with a mathematical model to generate a numerical index^1^ that correlates with other motor unit number estimation (MUNE) methods with faster or comparable acquisition times^2^. It is a feasible technique for multicentre research^3^ and recent guidelines have been proposed for consistent recording and analysis across laboratories^4^.

MUNIX is a candidate biomarker of longitudinal lower motor neuron (LMN) dysfunction with excellent reliability in limb muscles and potential utility in diseases such as amyotrophic lateral sclerosis (ALS) and peripheral neuropathies^2^. However, MUNIX of brainstem-innervated muscles remains technically challenging, which limits its application in conditions with diffuse LMN pathology. In ALS, MUNIX of facial nerve-innervated muscles has been explored as a potential correlate for bulbar dysfunction; but there was poor discrimination between scores from patients with bulbar weakness and controls^5,6^.

The upper trapezius is an alternative for probing LMNs from the bulbar region as it is innervated by the spinal accessory nerve (SAN) comprised of cranial and spinal roots^7^. Neurogenic denervation of the trapezius was demonstrated by needle EMG in patients with ALS and early bulbar weakness^8^. MUNIX of the upper trapezius is feasible and has lower mean values in ALS patients compared to healthy controls as demonstrated in two small studies^5,9^. Another study applied MUNIX from the upper trapezius in patients with spinal muscular atrophy (SMA) as a biomarker of LMN pathology^10^. Although adequate intra-rater reliability was recently reported in healthy volunteers^11^, there are no data available on inter-rater reliability. Here, we conducted a systematic review and meta-analysis of published MUNIX upper trapezius studies and assessed the inter- and intra-rater reliability of this parameter in twenty participants without peripheral nerve disorders.

## Methods

### Participants

We enrolled volunteers aged 18 years and older without a medical history or exposure to medications associated with a risk of neuropathy. The study was approved by the local ethics board and written informed consent was obtained from all subjects.

### Systematic review and meta-analysis

Medline and EMBASE were searched in February 2021 using the terms “MUNIX” AND (“trapezius” OR “spinal accessory nerve”) without language or date filters. Inclusion criteria were original articles reporting MUNIX in the upper trapezius of healthy volunteers. Two independent reviewers screened and extracted data from eligible articles.

### Procedures

Participants were seated upright with the active electrode (E1) placed over the motor point of the upper trapezius (about mid-point from C7 to acromion). The reference electrode was placed over the contralateral acromion to ensure isoelectrical recording and to avoid volume-conducted potentials from ipsilateral co-activated muscles. We stimulated the right SAN at the posterior sternocleidomastoid site, making all the efforts to achieve supramaximal CMAP, including required changes in E1 positioning^4^. CMAP baseline-peak amplitudes were recorded (filters 2Hz – 10kHz). Twenty 500-millisecond SIP epochs were recorded at five increasing levels of voluntary shoulder shrug (filters 20Hz – 10kHz). CMAP and MUNIX values were obtained from Natus VikingQuest™ with MUNIX software v22.

MUNIX sessions were performed 30 minutes apart by two blinded raters with all electrodes and markings removed from the volunteers in between tests. Two sessions were performed by a novice neurophysiology fellow (rater-1) and one session by a board-certified neurophysiologist with prior MUNIX experience (rater-2).

### Statistical Analyses

Mean MUNIX values, standard deviation (SD), and pooled 95% confidence interval (95%CI) of the mean were reported. Inter- and intra-rater reproducibility was calculated using two-way random effects model with multiple raters intraclass correlation (ICC [2,k]). The coefficient of variability (CoV) was calculated as the standard deviation over the mean with 95%CI through 10,000 bootstrap resamples with replacement. Rater-1 test-retest data were used to calculate intra-rater reproducibility, whereas rater-1 test and rater-2 test data were used for inter-rater reproducibility. Bland-Altman plots demonstrated intra- and inter-rater agreement. A sample size of 10 participants with 2 raters provided 83% power to detect an inter-rater ICC of 0.85 and null hypothesis ICC of 0.2 at two-sided alpha 0.05. A random-effects meta-analysis of single-group MUNIX means was estimated using the inverse variance method and DerSimonian-Laird τ^2^ estimator. Analyses were computed using SAS Studio® and R (metafor package).

## Results

Twenty volunteers (10 females) with mean age of 33 years (SD 9, range 23-56) were enrolled. Four published studies reported MUNIX in the upper trapezius for a total of 203 healthy volunteers (Figure A.1). In the enrolled sample, mean MUNIX was 209.6 (SD 78) without a floor or ceiling effect, while the pooled mean MUNIX was 176.2 (95%CI 155.7; 196.7) in a meta-analysis of all available data (Figure 1). Mean CMAP baseline-to-peak amplitude was 9.5mV (SD 2.6) in our sample.

**Figure 1.**
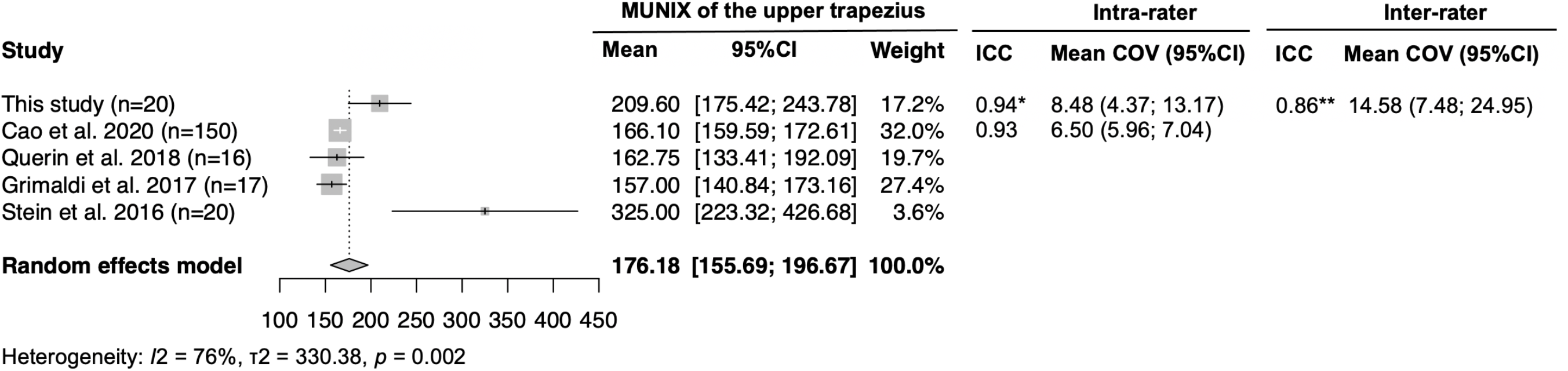
Forest plot of mean MUNIX of the upper trapezius in healthy volunteers from four published studies, in addition to data from this study. 95%CI of mean CoV for Cao et al. was estimated using the Mean ± (SD/√n x 1.96) from their published data. * n=20; ** n=10.

Inter-rater ICC (n=10) of 0.86 indicated adequate reliability between a novice and an experienced rater in this study. Intra-rater reliability (n=20) performed by the novice rater revealed excellent agreement, similar to the data reported by Cao et al.^11^ (Figure 1). Conforming to MUNIX guidelines, the inter- and intra-rater CoVs were below 20% with excellent agreement among raters as indicated by the Bland-Altman plots (Figure 2).

**Figure 2.**
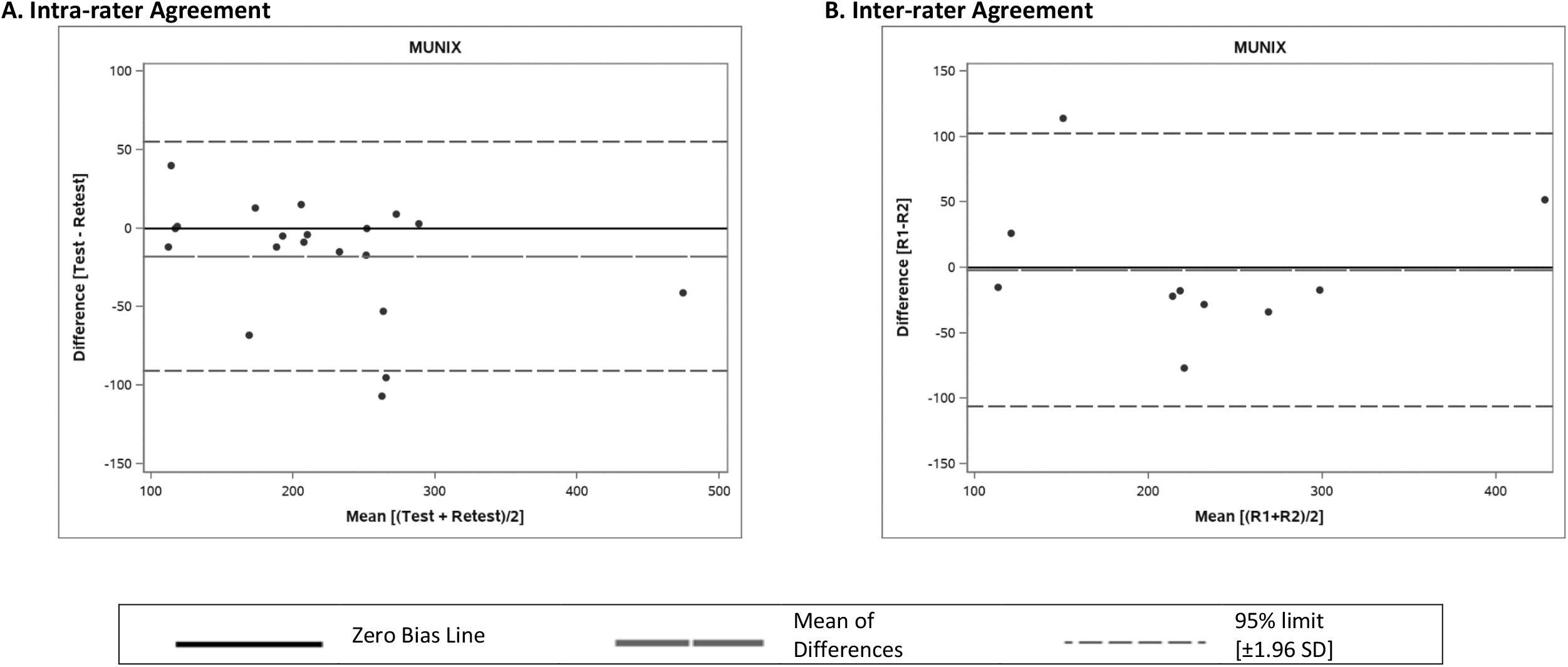
Bland Altman plots of intra- and inter-rater agreement. (A) Intra-rater agreement of MUNIX scores from Rater 1 test and re-test sessions. (B) Inter-rater agreement of MUNIX scores from Rater 1 test and Rater 2 test sessions.

There was significant heterogeneity in mean MUNIX values (I^2^ 76%), along with variability in age and sampling technique across studies (Table A.1). Mean age varied across studies (Figure A.2) and moderated mean MUNIX values in a meta-regression model (Figure A.3). Stein et al. reported significantly higher mean value (325) and larger SD (232) when applying a modified SIP recording technique in a relatively younger sample.

## Discussion

To serve as a global biomarker of functional motor units in diseases with diffuse LMN involvement, MUNIX must probe not only the cervical and lumbosacral-innervated regions, but also bulbar dysfunction. MUNIX of the upper trapezius can potentially correlate with brainstem LMN function and has been tested in early studies in ALS and SMA^5,9,10^. However, reliability data are incomplete for this measure, which limits its interpretation and implementation in multicentre studies. Here, we synthesized the published data on MUNIX upper trapezius in healthy controls and demonstrated excellent inter- and intra-rater reliability of this MUNIX parameter in otherwise healthy participants.

MUNIX recorded at the upper trapezius is a promising option relative to facial nerve-innervated muscles. It is less susceptible to a floor effect as the supramaximal CMAP recorded at the trapezius – a key component for MUNIX calculation – is larger compared to CMAP captured in facial muscles. The CMAP from trapezius also typically begins negative, while facial motor responses are often initially positive, making it less certain as to the origin of the potentials. Moreover, in ALS, trapezius voluntary activation against gravity is relatively preserved until advanced stages, allowing for feasible SIP recording as the disease progresses. It can also be performed with patients seated upright and wearing facial masks, avoiding the need for wheelchair transfers or interruption of non-invasive ventilation.

Following the guidelines for MUNIX acquisition^4^, recordings at the upper trapezius had reliable test-retest performance between a novice and senior MUNIX rater. This is encouraging for its use in multicentre studies where varying MUNIX skills are expected^3^. Test-retest reproducibility was excellent when performed by a novice rater in our sample, further supporting studies with longitudinal measures as previously reported by Cao et al^11^. While the inter- and intra-rater ICCs from healthy controls represent intrinsic technical variability, future studies should reassess the upper trapezius MUNIX reproducibility in disease scenarios to account for potential patient-level factors on reliability. Nevertheless, in limb muscles, MUNIX ICCs were comparable between controls and ALS patients^12^.

MUNIX is susceptible to variability in SIP sampling, particularly at low contraction levels^13^. This may partially account for the heterogeneity across studies and outlier mean values reported by Stein et al^9^. These authors pursued a continuous SIP sampling method at gradually increasing voluntary muscle contractions with post-hoc extraction of shorter-length epochs, in contrast to the recommended stepwise SIP recording protocol. Also, differences in mean age across studies, but not sex, contributed to heterogeneity and a younger sample was enrolled by Stein et al. Supporting this, age inversely correlated with MUNIX trapezius values in the large cohort reported by Cao et al^11^.

The most promising utility of MUNIX trapezius is as a bulbar LMN degeneration measure in ALS. While cross-sectional MUNIX comparisons could discriminate patients with ALS from controls^5,9^, longitudinal construct validity studies correlating MUNIX trapezius and clinical measures of speech and swallowing in patients with and without bulbar LMN dysfunction are warranted. Advanced speech assessment tools, such as speech-and-pause analysis of spoken passage recordings^14^, can also be used as anchors of bulbar function to correlate with MUNIX over time.

This study has limitations. The small number of MUNIX upper trapezius studies reduces the precision of the pooled estimates and meta-regression inferences. The generalizability of our reliability data is limited given the small and younger sample from a single site, despite our intra-rater ICC yielding similar to previously reported.

In conclusion, MUNIX upper trapezius is a reliable measure with excellent inter- and intra-rater reproducibility. Variability in MUNIX sampling techniques and age group of healthy controls account for heterogeneity in the mean MUNIX values. With further longitudinal construct and pharmacological surrogacy validation, this measure can serve as a biomarker of brainstem LMN function over time in diseases such as ALS.

## Data Availability

Data referred to in this manuscript will be made available upon request to the corresponding author.

## Acknowledgements

The authors thank Dr. Aude-Marie Grapperon and collaborators for sharing additional data of MUNIX of the upper trapezius in healthy controls from the Grimaldi et al. study.

## Funding Source

This study was funded by the generosity of philanthropic gifts to the Sunnybrook Foundation and our ALS research program by the Temerty family.

**Appendix A. Supplementary Material**

