## Supplementary Figures and Tables for "Motor Unit Number Index (MUNIX) of the Upper Trapezius: Reliability and Meta-Analysis"

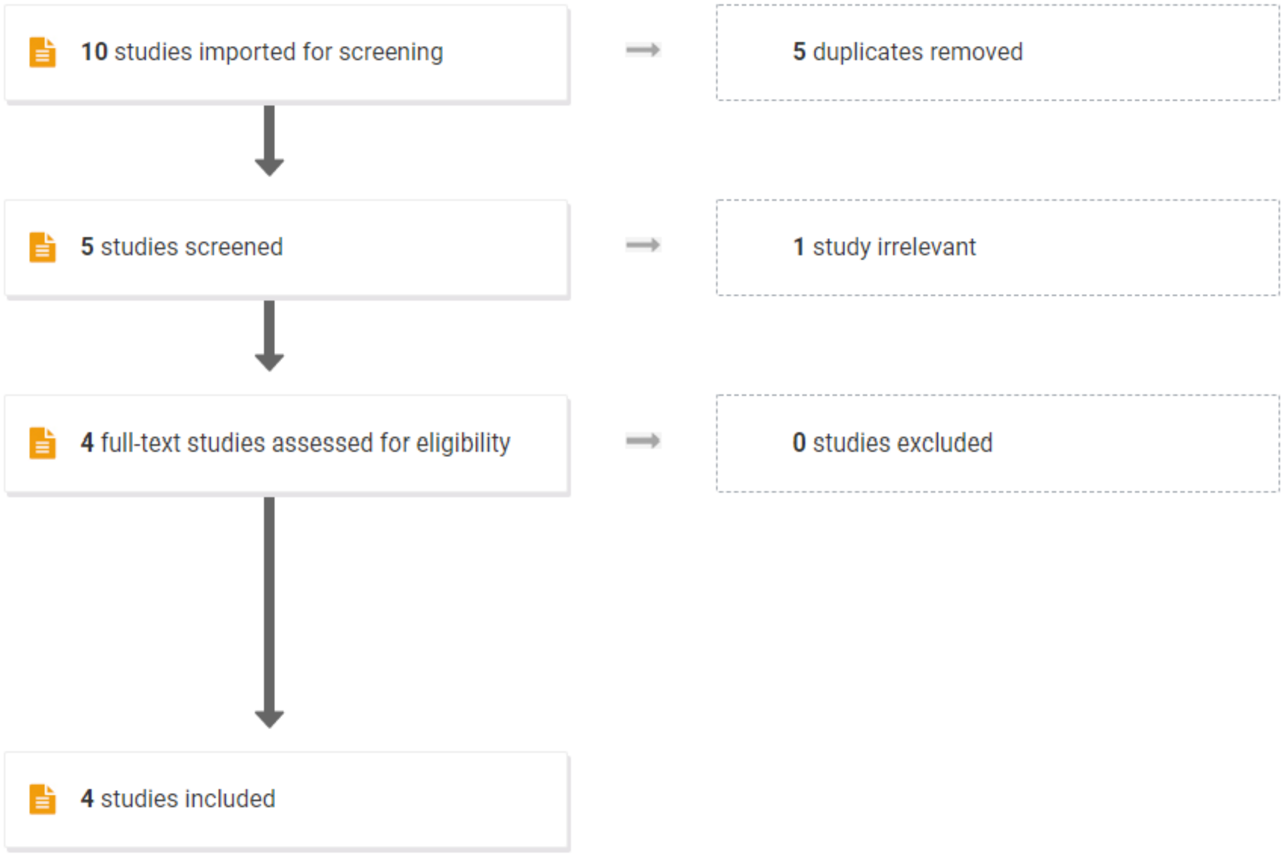


**Figure A.1.** PRISMA flow chart of articles selected for meta-analysis.

| *Study* | *n* | *Mean age (SD)* | *Females (%)* | *Hardware* | *Software* | *Technique* | *E1 repositioning for supramaximal CMAP?* | *Location of E2* | *Total number of SIPs* | *SIP epoch length* | *Levels of muscle contracti on* |
|---|---|---|---|---|---|---|---|---|---|---|---|
| ***This study*** | 20 | 33 (9) | 50 | Nicolet¨ VikingQuestª system (Natus) | Synergy v22 (Natus) | MUNIX guidelines (2018) | Yes | Contralateral acromion | 20 | 500 ms | 5 |
| ***Cao et al. 2020*** | 150 | 49 (17) | 49 | Nicolet EDx system (Natus) | Natus v22 | MUNIX guidelines (2018) | Yes | Not reported | At least 20 | 500 ms | 5 |
| ***Querin et al. 2018*** | 16 | 40 (13) | 38 | Nicolet¨ VikingQuestª system (Natus) | Natus v21 | Original MUNIX (2004) | Not reported | Not reported | At least 10 | 500 ms | Not reported |
| ***Grimaldi et al. 2017*** | 17 | 63 (7)* | 53 | Keypoint system (Medtronic) | Excel file provided by Keypoint | Original MUNIX (2004) | Yes | Ipsilateral acromion | 10 | 300 ms | 5 |
| ***Stein et al. 2016*** | 20 | 24 (3)* | 65 | Schwarzer topas system (Natus) | LabView Ver11 (National Instruments) | Continuous SIP | Yes | Not reported | 50 | Continuous | Not reported |

*Estimated SD

**Table A.1.** Demographics and qualitative summary of MUNIX techniques used among studies.

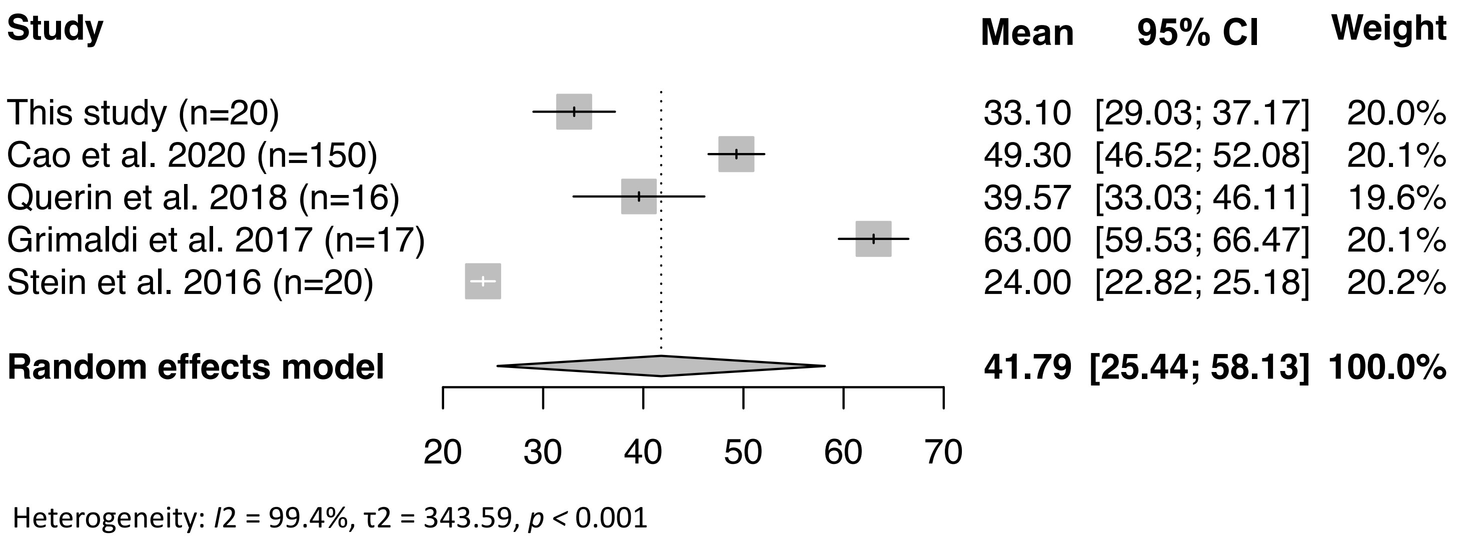


**Figure A.2a.** Ages varied significantly in between studies. This meta-analysis was run following the methodology in main manuscript.

| Study | Events | Total | Proportion | 95%–CI | Weight |
|---|---|---|---|---|---|
| This study (n=20) | 10 | 20 | 0.50 | [0.27; 0.73] | 9.1% |
| Cao et al. 2020 (n=150) | 74 | 150 | 0.49 | [0.41; 0.58] | 68.1% |
| Querin et al. 2018 (n=16) | 6 | 16 | 0.38 | [0.15; 0.65] | 6.8% |
| Grimaldi et al. 2017 (n=17) | 9 | 17 | 0.53 | [0.28; 0.77] | 7.7% |
| Stein et al. 2016 (n=20) | 13 | 20 | 0.65 | [0.41; 0.85] | 8.3% |
| **Random effects model** | | **223** | **0.50** | **[0.44; 0.57]** | **100.0%** |

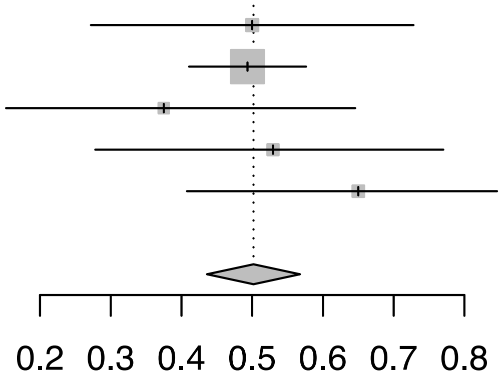


Heterogeneity: *I*2 = 0%, τ2 = 0.0, *p* = 0.59

**Figure A.2b.** Fairly balanced proportion of females and males was seen across studies. This random-effects meta-analysis of single-group proportions of females was run in R (meta package), using logit transformation of the data, inverse variance method and restricted maximum-likelihood estimator for $\tau^2$.

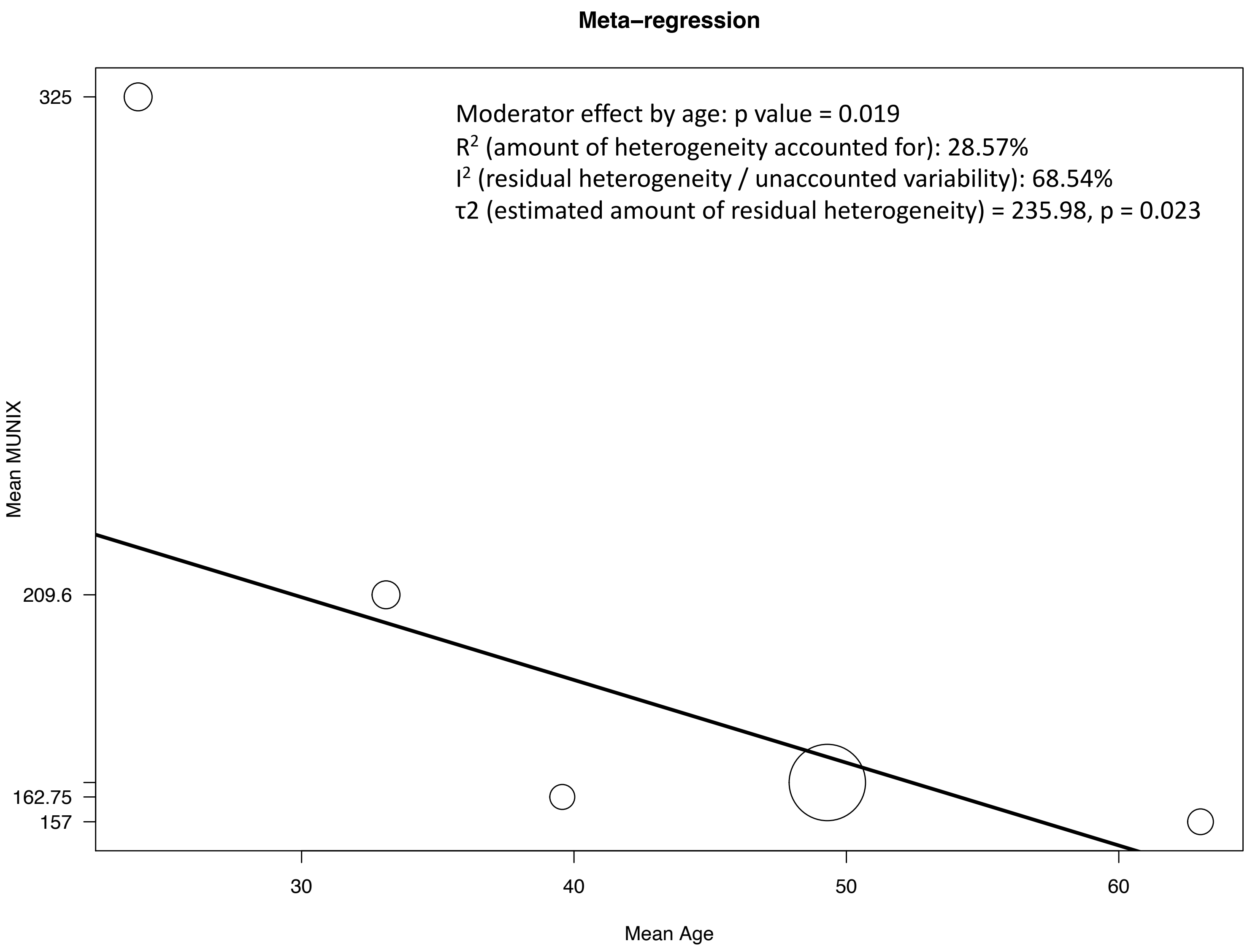


**Figure A.3.** Mean MUNIX of the upper trapezius was significantly moderated by mean age across four published studies, in addition to data from this study. Mixed-effect meta-regression model was fitted with DerSimonian-Laird estimator for $\tau^2$.
